# Care Across Contexts: Patterns of Caregiver-Infant Engagement in Spanish- and English-Speaking Families of Preterm Infants

**DOI:** 10.64898/2026.03.18.26348751

**Authors:** Pamela M. Rios, Virginia A. Marchman, Nuria L. Ontiveros Perez, Katherine E. Travis, Molly F. Lazarus, Melissa Scala, Heidi M. Feldman

## Abstract

**Objective:** To examine group differences and continuity in caregiving environments of infants born preterm from Spanish- and English-speaking families.

**Study Design:** We conducted a prospective cohort study of Spanish- (*n* = 17) and English-speaking (*n* = 23) families of infants born preterm (< 32 weeks gestation). Caregiver-infant engagement was assessed neonatally via hospital visitation and skin-to-skin (STS) care, and at home via child-directed adult word counts/hour (CD-AWC/hour) from all-day audio recordings.

**Result:** No significant group differences were observed in family visitation, neonatal STS care, or in-home verbal engagement, although STS care rates varied considerably, especially within Spanish-speaking families. Across both groups, greater STS care was associated with higher CD-AWC/hour at home.

**Conclusion:** Spanish- and English-speaking families showed comparable patterns of caregiver-infant engagement, as a group, however, many Spanish-speaking families engaged in less STS than English-speaking families. STS care predicted caregiver-infant verbal engagement at home, highlighting continuity from hospital to home.

---

A growing body of literature documents that children from Spanish-speaking families in the United States experience different environmental circumstances during infancy and toddlerhood than do children from English-speaking families. In California and elsewhere, for example, Latinx Spanish-speaking families are disproportionately represented among low-income households^1^, which is associated with limited resources and access to early social support systems, which in turn shape caregiving opportunities. Moreover, research has identified differences in parenting beliefs and practices between Hispanic, Spanish-speaking families and non-Hispanic, English-speaking families, reflecting socioculturally-patterned approaches to early caregiving.^2^ Infants in Spanish- and English-speaking households may therefore experience different early caregiving contexts that are shaped by the intersection of socioeconomic factors and distinctive cultural practices.

These environmental and sociocultural factors may be particularly consequential for families of infants born preterm, who require prolonged and complex interactions with healthcare systems and workers. For both Spanish- and English-speaking families, such interactions substantially shape the circumstances experienced in the hospital. Spanish-speaking caregivers, in particular, may face sociocultural barriers, including limited comprehension of verbal and written information, reduced interpreter access, and low levels of institutional support.^4–6^ Low socioeconomic status (SES), which is also associated with elevated risks of preterm birth^3^, may further compound these challenges and affect families’ ability to meaningfully engage in care.

We begin this exploration of caregiver-infant engagement in the neonatal period. The neonatal intensive care unit (NICU) plays a critical role in medically stabilizing infants born preterm, but it also represents an early context in which caregivers can begin engaging in developmentally-relevant interactions. Families are encouraged to visit their child in the NICU, but visitation rates may vary due to psychosocial, structural, and socioeconomic factors.^7–9^ Within this early and often stressful caregiving context, developmental care practices such as skin-to-skin (STS) care, in which a caregiver holds their infant against their bare chest, have emerged as a critical form of caregiver-infant engagement associated with improved neurodevelopmental outcomes among children born preterm.^10,11^ Prior work has documented disparities in STS care rates, showing that infants whose families speak a language other than English or come from lower-SES backgrounds experience less STS care than infants of English-speaking or higher-SES families.^12^ Rates of visitation and STS during hospitalization are objective indices of variation in caregiver-infant engagement among families of preterm infants and provide a window into variability of caregiving that may carry over into the home environment.

Moreover, emerging work suggests that when infants born preterm are discharged, levels of STS care established in the hospital may influence or predict levels of caregiver-infant verbal engagement at home.^16^ Despite documented differences in rates of STS care as a function of language group, few studies have asked whether patterns of continuity from hospital to home vary in linguistically-diverse households. Examining caregiving practices in families from different language backgrounds may help clarify sociolinguistic factors that shape patterns of caregiver-infant engagement.

The current descriptive cohort study of infants born preterm from Spanish- and English-speaking families considered aspects of the caregiving environment in both the hospital and home contexts. Specifically, we compared Spanish- and English-speaking families on three objective measures of caregiver-infant engagement: rates of hospital visitation, rates of neonatal STS care, and amount of caregiver-infant verbal engagement at home. Based on prior studies, it was hypothesized that we would see differences in these three indices of the caregiving environment related to the broader sociocultural characteristics of households, including variation in linguistically-concordant healthcare communication, institutional support, and culturally-patterned beliefs about caregiver-infant interactions. In this study, language group was conceptualized as a marker of these broader contextual dynamics.

This study also investigated the extent to which rates of engagement in the hospital were associated with levels of caregiver-infant verbal engagement at home. Our main focus was whether any evidence of continuity from hospital to home was moderated by language group.

Critically, language-group differences were examined controlling for SES, a known confound with language status in the Spanish-speaking population. The findings from this study may aid in clarifying whether and how sociocultural factors influence caregiving practices and inform the implementation of interventions that effectively support diverse families of preterm infants.

## Methods

### Participants

Participants (*n* = 40) were infants born very preterm, prospectively recruited between August 2020 and September 2024 from Lucile Packard Children’s Hospital (LPCH) at Stanford and California’s High Risk Infant Follow-Up Clinic (HRIF). Infants were included if they were born < 32 weeks gestational age (GA), had no congenital or genetic anomalies, no hearing or vision loss, had caregivers who primarily spoke English or Spanish, were inborn with a hospital stay length > 20 days, and had caregiver-infant verbal engagement data available at either 9 (*n* = 18) or 15 months (*n* = 22). An additional *n* = 27 children were considered for analysis but were excluded because they were outborn (*n* = 13), their hospital stay was shorter than 20 days (*n* = 1), or home engagement data were missing (*n* = 13). This study was reviewed and approved by the Stanford School of Medicine Institutional Review Board (IRB; Protocol #53904, #44480), informed consent was obtained by the primary caregiver, and the study was performed in accordance with the Declaration of Helsinki.

### Clinical and Demographic Information

Demographic and clinical characteristics of infants, including GA (days), length of hospital stay (days), and child sex assigned at birth (male or female), were extracted from the electronic medical record (EMR). Race and ethnicity were reported by caregivers at the time of enrollment. Additionally, caregivers completed a detailed language background questionnaire asking the number of hours per day each caregiver typically spoke each language to the child, estimates that were then converted to percentages. Families were classified as English speaking if the child was reported to be exposed to ≥ 60% English or if both caregivers’ native language was English. Families were classified as Spanish speaking if it was reported that the child was exposed to ≥ 60% Spanish or if one or both caregivers reported that Spanish or Spanish/English was their native language. Socioeconomic status (SES) was measured using an updated version of the Hollingshead Four Factor Index^17^, which combines the education and occupation of both caregivers into a single composite score (range = 8 to 66). Because health conditions could influence rates of STS care, clinical data regarding four common comorbidities of premature birth were also obtained from the EMR. The co-morbidities were: bronchopulmonary dysplasia (BPD), defined as treatment with supplemental oxygen at 36 weeks postmenstrual age^18^; intraventricular hemorrhage (IVH) by any grade present; sepsis by positive blood culture or > seven days of antibiotics; and necrotizing enterocolitis by medical or surgical diagnosis. From these conditions, a binary health acuity variable was derived, classifying infants as having either none (health acuity = 0) or one or more (health acuity = 1) of these conditions.

### Measures

#### Family Visitation and Skin-to-Skin Care in the Hospital

At Lucile Packard Children’s Hospital (LPCH), nurses routinely document all instances of family visitation to bedside. To account for varying lengths of infant hospitalization, family visitation (visits/day) was computed as the total number of visitation instances divided by the number of days of infant hospital stay. Bedside nurses routinely document all instances of STS care, the approximate duration of each session in minutes, and the individual(s) involved. Rates of family-delivered STS care (STS minutes/day) were calculated as total minutes of STS care provided by one or more family members divided by the number of days of infant hospital stay.

#### Caregiver-Infant Verbal Engagement in the Home

Caregiver-infant verbal engagement in the home was assessed using day-long audio recordings via the Language Environment Analyses (LENA) system.^19^ Analyses used recordings collected when the child was either 9- or 15-months of age. Recordings at older ages were prioritized such that the 15-month recording was used if available, and if not, the recording collected at approximately child age 9-months was analyzed. The LENA system consists of a high-quality digital recording device worn by infants in specialized clothing. At all time points, families were instructed to begin recording during a typical day at home and to allow the LENA recorder to run automatically for up to 16 hours. The LENA speech-recognition software later classifies all adult speech that is “near and clear” to the child and generates adult word counts (AWC) for each 5-minute segment of the recording. Recordings were subsequently cleaned using an automated classifier developed for English- and Spanish-speaking families to identify those 5-minute segments during which the child was not likely to be sleeping and that were most likely to contain child-directed speech (CDS).^20^ Child-directed adult word counts/hour (CD-AWC/hour) were computed as the sum of AWC across the non-sleep, child-directed segments, divided by the length of the cleaned recording. Consistent with prior studies that have documented that AWC is stable in this period^19^, we found that CD-AWC/hour was strongly correlated in those families for whom recordings were assessed at both 9 and 15 months, *r(14)* = 0.79, *p <* 0.001.

### Statistical Analyses

All analyses were conducted in R version 4.5.2. Hierarchical linear mixed-effects models^21^ examined language group differences in family visitation, STS care rates, and CD-AWC/hour. A power analysis indicated that our sample size provides > .8 power to detect a large effect (> .4) of language group. We then tested predictive relations between family visitation and STS care rates and later CD-AWC/hour. Inclusion of the interaction terms of language group by visitation and language group by STS rate tested whether relations to later CD-AWC/hour were moderated by language group. A power analysis indicated that our sample size was sufficiently powered (> .8) to detect a *Δr²* of .20, a medium effect. All models included family ID as a random intercept to account for clustering among families with multiples. SES was selected *a priori* as a covariate, given its anticipated relevance to caregiver engagement measures. Given the difference in GA by language group, GA was also included as a covariate. The examination of effects of STS care rates also controlling for family visitation distinguished the effects of engaging in STS care from general patterns of hospital visitation.

## Results

### Language Group Differences during the Neonatal Hospital Stay

Participants (*n* = 40) were 17 Spanish-speaking and 23 English-speaking families of infants born very preterm. About half of the infants were female in both groups (Spanish speaking: 47.1%; English speaking: 47.8%) reflecting no significant difference in sex distribution, χ²(1) = 0.00, *p* = 1.00. Infants from Spanish-speaking families were hospitalized for an average of 82.1 days (SD = 50.7), while infants from English-speaking families were hospitalized for 62.5 days (SD = 32.4); this difference was not statistically significant, *t*(38) = - 1.50, *p* = 0.15. Infants were also matched in health acuity across groups, with a total of 11 Spanish-speaking infants (64.7%) and 12 English-speaking infants (52.2%), χ²(1) = 0.22, *p* = 0.64, having one or more common comorbidities of premature birth. Moreover, individual preliminary linear mixed-effects models indicated that health acuity was not significantly associated with family visitation, β = -0.13, SE = 0.19, *p* = 0.51, STS care rates, β = -12.12, SE = 9.92, *p* = 0.23, nor CD-AWC/hour at home, β = -43.41, SE = 241.11, *p* = 0.86. All of the families in the Spanish-speaking group reported child ethnicity as Hispanic (100%), whereas 5 families (22%) in the English-speaking group did so. Mean GA at birth (measured in days) differed significantly by language group; infants in the Spanish-speaking group were born earlier than those in the English-speaking group (Spanish M = 196.4 days, SD = 19.5, English M = 206.6 days, SD = 12.4), *t*(38) = 2.03, *p* = 0.05. As predicted, Spanish-speaking participants reported significantly lower SES than English-speaking families (Spanish M = 22.5, SD = 10.5, English M = 54.7, SD = 8.5), *t*(38) = 10.76, *p* < 0.001. Both GA at birth and SES were included as covariates in all primary analyses.

Table 1 presents the adjusted linear mixed-effects models examining differences in caregiver-infant engagement by language group, with Figure 1 illustrating the corresponding model-estimated values. Across language groups, family visitation was approximately 1, corresponding to about one hospital visit per day, on average. Visitation rates were comparable in Spanish- and English-speaking families, β = 0.03, SE = 0.44, *p* = 0.94.

**Figure 1.**
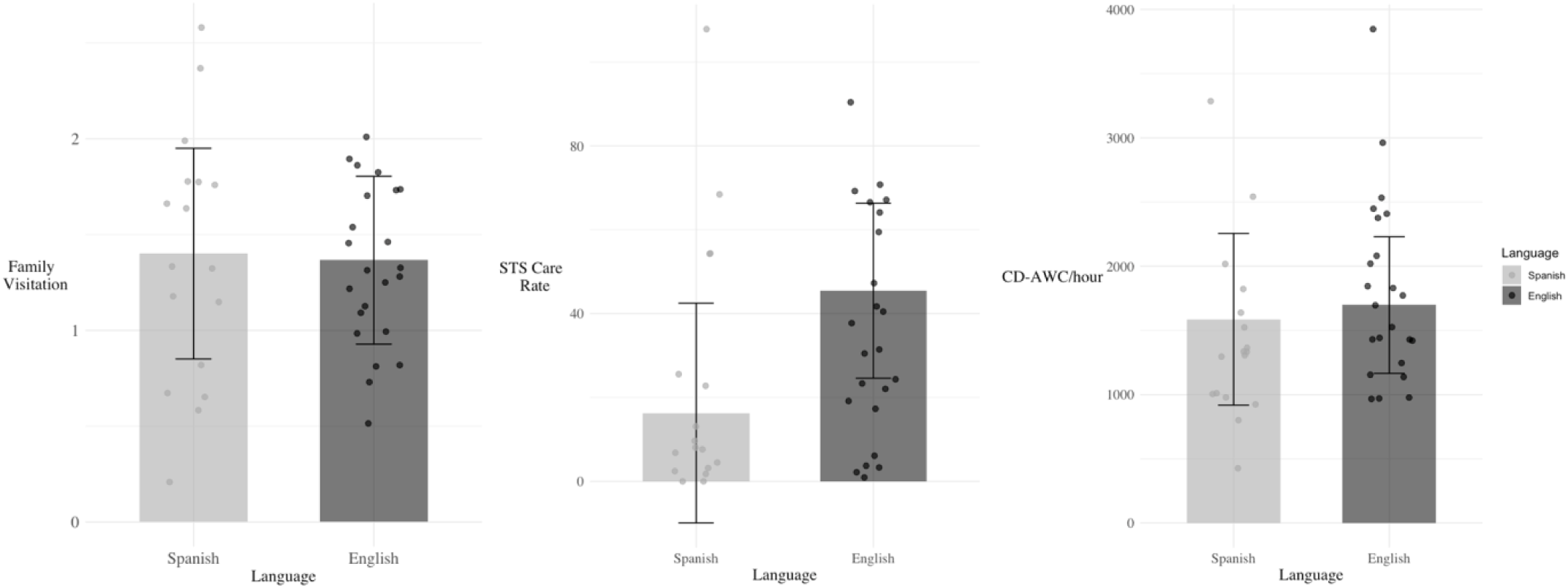
Estimated marginal means of family visitation, STS care rates, and CD-AWC/hour by language group, controlling for relevant covariates. *Note:* Error bars indicate standard errors. Points represent individual observations. Models include random intercepts of family ID to account for twins/triplets.

**Table 1.**
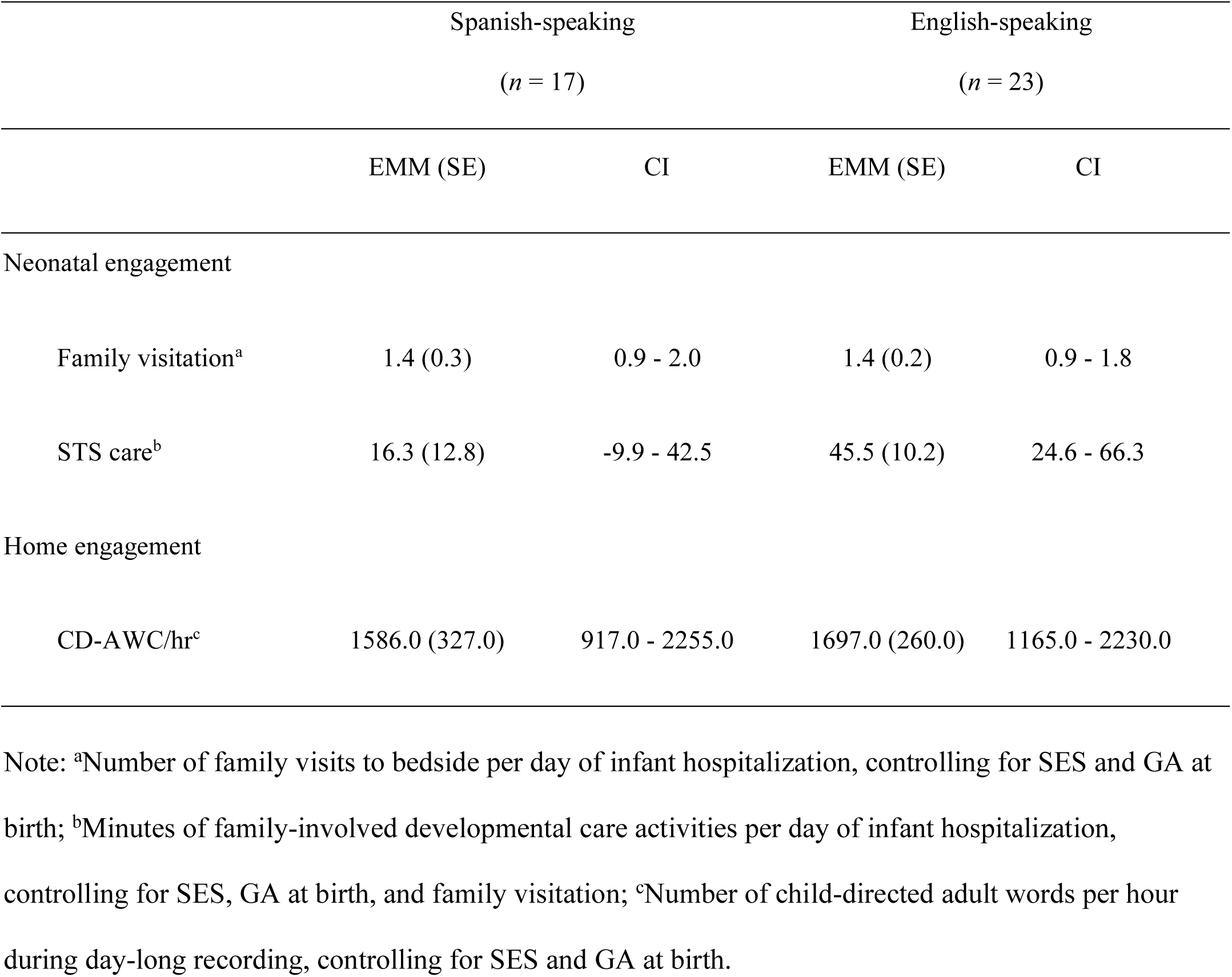
Estimated marginal means for neonatal and home engagement variables for infants by language group, controlling for relevant covariates (total *n* = 40).

| | Spanish-speaking<br>($n = 17$) | | English-speaking<br>($n = 23$) | |
| --- | --- | --- | --- | --- |
|  | EMM (SE) | CI | EMM (SE) | CI |
| Neonatal engagement |  |  |  |  |
| Family visitation<sup>a</sup> | 1.4 (0.3) | 0.9 - 2.0 | 1.4 (0.2) | 0.9 - 1.8 |
| STS care<sup>b</sup> | 16.3 (12.8) | -9.9 - 42.5 | 45.5 (10.2) | 24.6 - 66.3 |
| Home engagement |  |  |  |  |
| CD-AWC/hr<sup>c</sup> | 1586.0 (327.0) | 917.0 - 2255.0 | 1697.0 (260.0) | 1165.0 - 2230.0 |

Note: <sup>a</sup>Number of family visits to bedside per day of infant hospitalization, controlling for SES and GA at birth; <sup>b</sup>Minutes of family-involved developmental care activities per day of infant hospitalization, controlling for SES, GA at birth, and family visitation; <sup>c</sup>Number of child-directed adult words per hour during day-long recording, controlling for SES and GA at birth.

Across language groups, STS care rates averaged approximately 30 minutes per day with substantial individual variability, from no STS care at all to 108 minutes per day. When controlling for SES and GA at birth, the difference between Spanish- and English-speaking families was not statistically significant, β = -28.67, SE = 22.11, *p* = 0.21. Since family visitation and STS care rates were positively correlated, indicating that families who visited their infants more often also tended to engage in more STS care, *r*(38) = 0.34, *p* = 0.03, we also examined group differences in STS care rates controlling for visitation. We found that language group differences remained non-significant, indicating no statistically significant difference between Spanish- and English-speaking families, as depicted in Table 1 and Figure 1, β = -29.18, SE = 20.92, *p* = 0.17. However, descriptively, while a few Spanish-speaking families showed STS care rates similar to English-speaking families, most Spanish-speaking families engaged in rates of STS care that fell in the bottom of the distribution compared to English-speaking families, as visible in Figure 1.

### Language Group Differences in the Home Environment

Total all-day home audio recording durations were about 15 hours, on average in both groups (Spanish M = 14.3, SD = 3.7; English M = 15.2, SD = 1.9), *t*(38) = 0.91, *p* = 0.37. Upon cleaning for non-sleep, child-directed segments, the average duration of recordings for Spanish-speaking families was 7.5 hours (SD = 3.2) and 7.0 hours (SD = 2.6) for English-speaking families, *t*(38) = -0.48, *p* = 0.63. Across both language groups, caregiver-infant verbal engagement at home averaged approximately 1700 CD-AWC/hour, with remarkably similar rates in the Spanish- and English-speaking families, β = -111.15, SE = 533.41, *p* = 0.84 (see Table 1 and Figure 1).

### Continuity from Hospital to Home

To examine continuity in caregiving across contexts, we next assessed whether infants from Spanish- and English-speaking homes with higher levels of verbal engagement at home were likely to have been visited more and/or received more STS care in the hospital (Table 2). Model 1 again showed that CD-AWC/hr in the home was not significantly different as a function of language group, controlling for SES and GA at birth; language group and SES together explained 5% of the variance in CD-AWC/hr. Model 2 added family visitation, which did not significantly improve model fit, accounting for only 3% additional variance, χ²(1) = 1.35, *p* = 0.24. Model 3 asked whether any effects of visitation were moderated by language group; the interaction term did not increase overall model fit, χ²(1) = 0.08, *p* = 0.78.

**Table 2.**
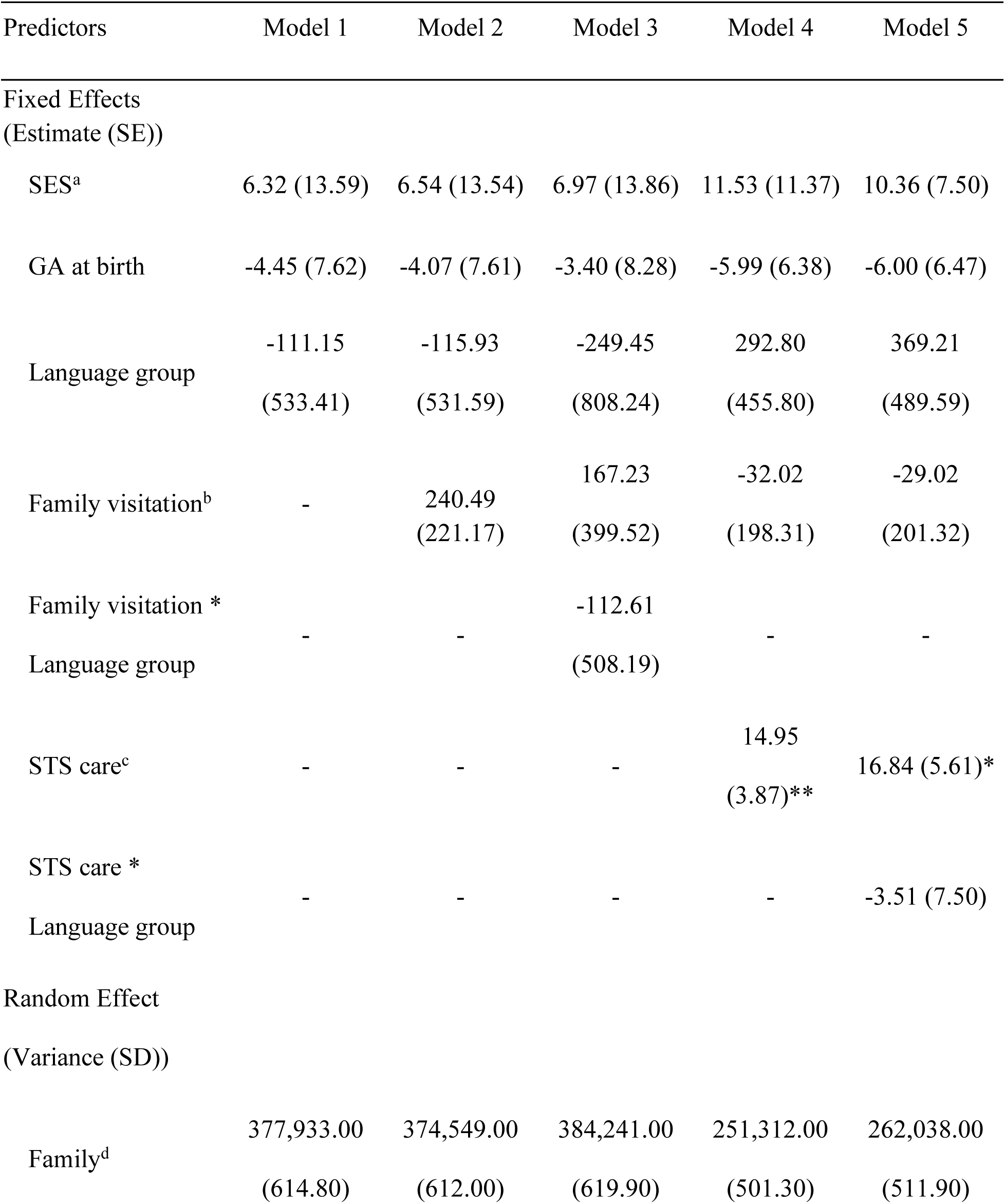
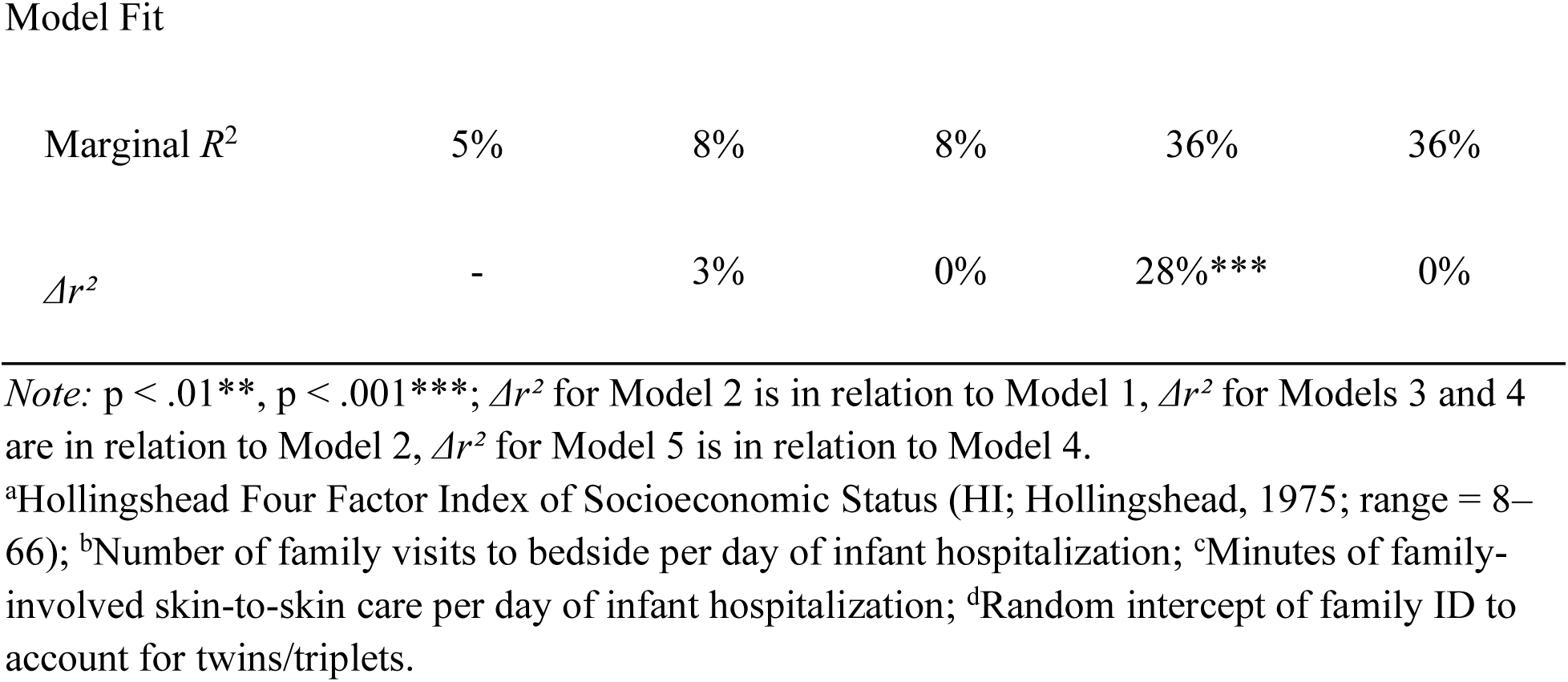
Linear mixed-effects models predicting CD-AWC/hour, after consideration of covariates, including random intercept for family (*n* = 40).

Next, we asked the degree to which STS care rates predicted CD-AWC/hr, controlling for SES, GA at birth, language group, and visitation rates (Model 4). STS care was a significant predictor of CD-AWC/hour at home, β = 14.95, SE = 3.87, *p* < 0.01, accounting for 28% additional variance and significantly improving model fit, χ²(1) = 14.64, *p* < 0.001. Each additional minute per day of skin-to-skin care during the infant’s hospital stay was associated with approximately 15 more CD-AWC/hour. Finally, Model 5 tested whether the relation between STS care rates and CD-AWC/hour differed by language groups; this interaction term did not significantly improve model fit, χ²(1) = 0.24, *p* = 0.62. Figure 2 illustrates the best-fitting model (Model 4), showing that infants who received more STS care in the neonatal period later lived in homes with higher levels of caregiver-infant verbal engagement, regardless of SES, gestational age, visitation rate, and language group.

**Figure 2.**
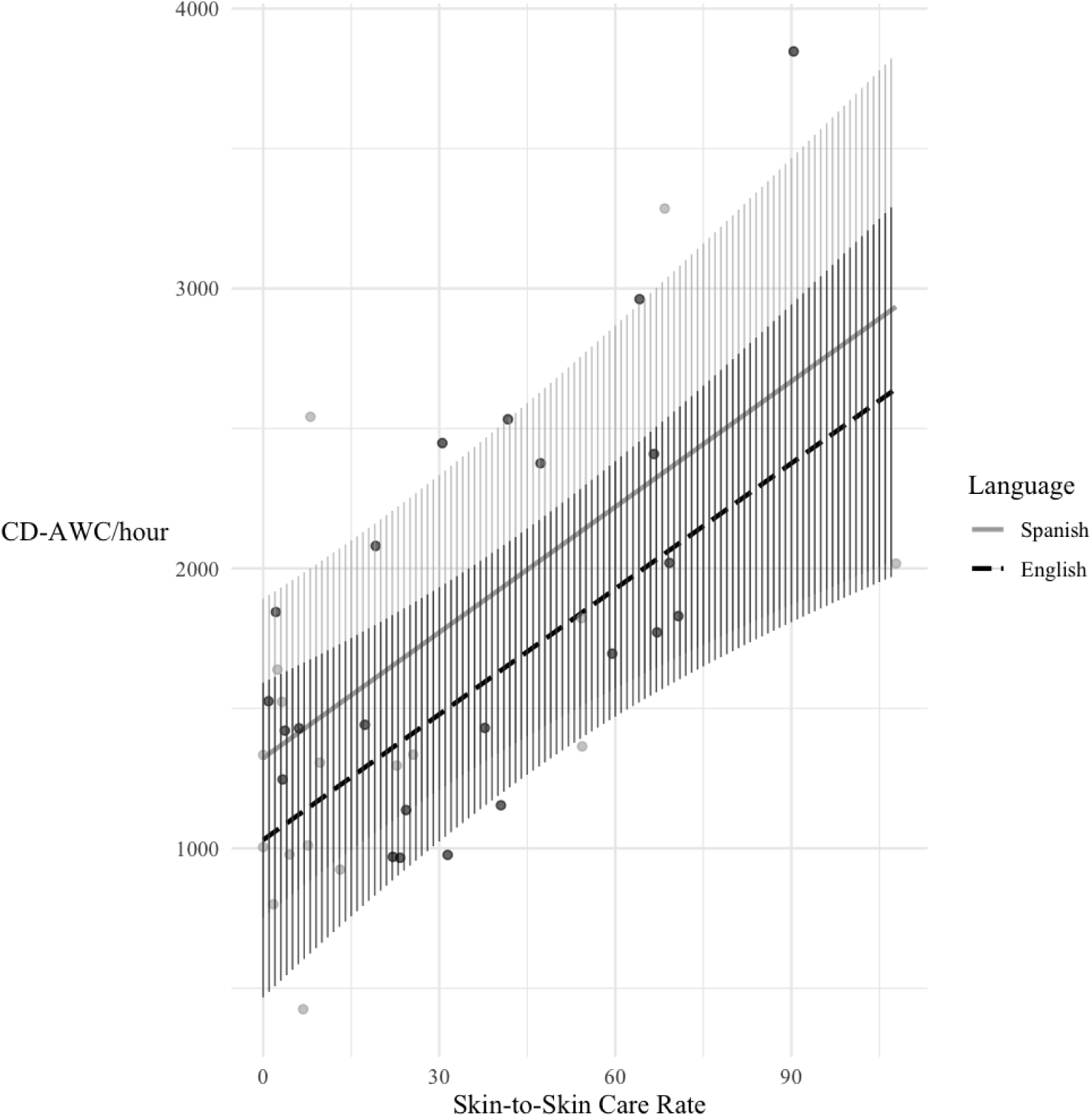
Model predictions showing relations between STS care rates in the NICU and CD-AWC/hour in the home by language group, controlling for SES, GA at birth, and family visitation. *Note:* Lines represent model-predicted values for English- and Spanish-speaking caregivers. Hatched areas indicate 95% confidence intervals of the model predictions. Points represent individual observations. Model includes a random intercept of family ID to account for twins/triplets.

## Discussion

This study compared Spanish- and English-speaking families of preterm infants on rates of hospital- and home-based caregiver-infant engagement. We found that, across language group, families demonstrated comparable rates of both visitation and in-home caregiver-infant verbal engagement. Although prior research has documented that specific factors, including SES^12^, language group^12^, and patient-staff language concordance^8^ may influence rates of hospital visitation for infants born preterm, we found comparable rates. This finding suggests that, within this cohort, families had similar levels of access to the NICU environment and opportunities for caregiver presence throughout hospitalization. Such equivalence in visitation may reflect improvements in unit-level practices or supports that facilitate family presence, although these factors were not directly examined. Understanding the conditions under which equitable visitation is achieved may inform efforts to promote caregiver involvement during the neonatal period. The comparable levels of in-home engagement in our Spanish- and English-speaking families, on average, also suggests similarities in the daily experiences of infants in our sample, in spite of their different sociocultural backgrounds.

Prior work has also documented disparities in STS care among families of infants born preterm, with low-SES, non-English speakers experiencing less STS compared to high-SES, English-speaking families.^12^ In the present study, rates of STS showed trends in this direction but did not significantly differ between Spanish- and English-speaking families. The lack of statistical differences may reflect characteristics of our sample or features of the broader care context, such as unit-level practices that support family engagement and/or increased awareness of language- and SES-related disparities in neonatal care. Descriptively, however, most Spanish-speaking families engaged in lower STS than English-speaking families. This pattern may reflect sociocultural barriers that limit caregivers’ access to staff instruction regarding STS or opportunities for clarification and encouragement. Alternatively, it may reflect a tendency for low-income, Spanish-speaking families to visit during evenings or weekends, when nursing staff are less likely to facilitate or support STS. Future studies with larger samples are needed to determine whether these trends represent meaningful group differences and to identify the mechanisms that support or constrain early STS care engagement across language groups.

Additionally, greater engagement in STS care during the infant’s hospital stay was associated with higher levels of CD-AWC/hour in the home across families. These findings extend prior work demonstrating continuity in caregiving patterns from the hospital to the home.^16^ Importantly, language group did not moderate these effects. In both Spanish- and English-speaking families, greater engagement in STS care in the hospital, but not visitation rates, was associated with higher levels of caregiver-infant verbal engagement in the home. These findings suggest that the mechanisms linking early STS care to later caregiver-infant verbal engagement are robust across families from different language backgrounds. Thus, interventions to support STS care in the hospital are likely to benefit families regardless of language background, offering a clear target for promoting sustained caregiver-infant engagement. Future studies must explore whether interventions that begin in the neonatal period will be effective for promoting levels of caregiver-infant engagement later in development.

Strengths of this study include the careful classification of caregiver language, based on their own reports of language use at home, reducing ambiguity and supporting clear interpretations of language-related patterns. Another strength is the measure of STS care, based on nurse charting; nurses at LPCH undergo rigorous training and follow standardized procedures for documenting these early caregiving activities, reinforcing the accuracy of this measure.^22^

Additionally, our measure of caregiver-infant verbal engagement in the home environment was also objective, i,e., audio recordings without an observer present, which captures all speech directed to the child throughout the day. Although the CD-AWC/hour measure does not allow attribution of speech quantity to specific caregivers and cannot capture forms of non-verbal communication, it nonetheless offers a more objective, extensive assessment of language exposure in the home than other available methods.

This study had limitations. While the sample was diverse, it was of modest sample size, which may have limited our ability to detect language group differences or moderation effects. In addition, as a single site study, we cannot expand on how hospital- or population-level factors may influence measures of caregiver-infant engagement at other sites. Still, these findings provide valuable insights into caregiving patterns within this population and can establish a foundation for future multi-site research. Further research is also needed to examine how variation in hospital-based supports relates to engagement in early caregiving activities and whether these patterns generalize to lower-resourced clinical settings. Finally, our design is correlational and therefore, we cannot draw causal inferences.

In conclusion, this study provides insights into the degree to which Spanish- and English-speaking families of children born preterm differ in levels of early caregiver-infant engagement. Although differences in STS participation by language group were not statistically significant in this sample, observed patterns suggest that variation in levels of early engagement in STS care, in particular, warrants further investigation. Importantly, greater STS care in the hospital was associated with higher caregiver-infant verbal engagement at home, underscoring the continuity of interaction across hospital and home contexts. These findings highlight the neonatal hospitalization as a critical window for supporting caregiver-infant engagement and suggest that early interactive experiences may have lasting implications for the home language environment. Future work should examine whether structured, culturally-responsive interventions delivered during hospitalization can strengthen early engagement and sustain caregiver-infant interaction over time. A prospective clinical trial designed to enhance early STS and caregiver communication may help determine whether targeted NICU-based support can meaningfully influence longer-term developmental trajectories for infants from diverse contexts.

## Data Availability

Deidentified individual participant data will be made available upon request.

## Notes

**Conflict of Interest Disclosure:** The authors have no conflict of interest.

**Funding/Support:** This research work was supported by grants from the National Institutes of Health (H.M. Feldman, PI: 2R01-HD069150; K.E. Travis, PI:5R00-HD8474904) and the Stanford Maternal and Child Health Research Institute (K.E. Travis, Faculty Scholars Award).

### Competing Interest Statement

The authors have declared no competing interest.

### Funding Statement

This research work was supported by grants from the National Institutes of Health (H.M. Feldman, PI: 2R01-HD069150; K.E. Travis, PI:5R00-HD8474904) and the Stanford Maternal and Child Health Research Institute (K.E. Travis, Faculty Scholars Award).

### Author Declarations

The Institutional Review Board of the Stanford School of Medicine (Protocol #53904, #44480) gave ethical approval for this work.

## References

1. Poverty in California. Public Policy Institute of California. Accessed December 3, 2025. https://www.ppic.org/publication/poverty-in-california/

2. Keels M. Ethnic group differences in early head start parents’ parenting beliefs and practices and links to children’s early cognitive development. Early Child Res Q. 2009;24(4):381–397. doi:10.1016/j.ecresq.2009.08.002

3. Wild KT, Betancourt LM, Brodsky NL, Hurt H. The effect of socioeconomic status on the language outcome of preterm infants at toddler age. Early Hum Dev. 2013;89(9):743–746. doi:10.1016/j.earlhumdev.2013.05.008

4. Fisher CR, Bourque SL, Palau MA, Nino de Guzman Ramirez M, Hwang SS. NICU Caregiver Communication Preferences and Disparities by Primary Language: A Qualitative Study. Hosp Pediatr. 2024;14(11):937–944. doi:10.1542/hpeds.2024-007798

5. Zaylskie LE, Zickafoose JS, Leech AA, Jennings B, Curcio NM, Griffith KN. Health care access, utilization, and quality for children in English versus Spanish-speaking households. Health Aff Sch. 2025;3(3):qxaf039. doi:10.1093/haschl/qxaf039

6. Palau MA, Meier MR, Brinton JT, Hwang SS, Roosevelt GE, Parker TA. The impact of parental primary language on communication in the neonatal intensive care unit. J Perinatol. 2019;39(2):307–313. doi:10.1038/s41372-018-0295-4

7. Gonya J, Nelin LD. Factors associated with maternal visitation and participation in skin-to-skin care in an all referral level IIIc NICU. Acta Paediatr. 2013;102(2):e53–e56. doi:10.1111/apa.12064

8. Latva R, Lehtonen L, Salmelin RK, Tamminen T. Visits by the family to the neonatal intensive care unit. Acta Paediatr. 2007;96(2):215–220. doi:10.1111/j.1651-2227.2007.00053.x

9. Harris LM, Shabanova V, Martinez-Brockman JL, et al. Parent and grandparent neonatal intensive care unit visitation for preterm infants. J Perinatol. 2024;44(3):419–427. doi:10.1038/s41372-023-01745-x

10. Soleimani F, Azari N, Ghiasvand H, Shahrokhi A, Fatollahierad S. Do NICU developmental care improve cognitive and motor outcomes for preterm infants? A systematic review and meta-analysis. BMC Pediatr. 2020;20(1):67. doi:10.1186/s12887-020-1953-1

11. Lazarus MF, Marchman VA, Brignoni-Pérez E, et al. Inpatient Skin-to-skin Care Predicts 12-Month Neurodevelopmental Outcomes in Very Preterm Infants. J Pediatr. 2024;274. doi:10.1016/j.jpeds.2024.114190

12. Brignoni-Pérez E, Scala M, Feldman HM, Marchman VA, Travis KE. Disparities in Kangaroo Care for Premature Infants in the Neonatal Intensive Care Unit. J Dev Behav Pediatr. 2022;43(5):e304. doi:10.1097/DBP.0000000000001029

13. Loi EC, Vaca KEC, Ashland MD, Marchman VA, Fernald A, Feldman HM. Quality of caregiver-child play interactions with toddlers born preterm and full term: Antecedents and language outcome. Early Hum Dev. 2017;115:110–117. doi:10.1016/j.earlhumdev.2017.10.001

14. Adams KA, Marchman VA, Loi EC, Ashland MD, Fernald A, Feldman HM. Caregiver Talk and Medical Risk as Predictors of Language Outcomes in Full Term and Preterm Toddlers. Child Dev. 2018;89(5):1674–1690. doi:10.1111/cdev.12818

15. Dailey S, Bergelson E. Language input to infants of different socioeconomic statuses: A quantitative meta-analysis. Dev Sci. 2022;25(3):e13192. doi:10.1111/desc.13192

16. Rios PM, Marchman VA, Lazarus MF, et al. From Hospital to Home: Continuity between Skin-to-Skin Care and Later Verbal Engagement in Infants Born Preterm. medRxiv. Preprint posted online July 3, 2025:2025.07.02.25330762. doi:10.1101/2025.07.02.25330762

17. Hollingshead, A. B. (2011). Four Factor Index of Social Status. Yale Journal of Sociology, 8, 21–51. - References - Scientific Research Publishing. Accessed January 2, 2026. https://www.scirp.org/reference/referencespapers?referenceid=1581486

18. Jensen EA, Dysart K, Gantz MG, et al. The Diagnosis of Bronchopulmonary Dysplasia in Very Preterm Infants. An Evidence-based Approach. Am J Respir Crit Care Med. 2019;200(6):751–759. doi:10.1164/rccm.201812-2348OC

19. Gilkerson J, Richards JA, Warren SF, et al. Mapping the Early Language Environment Using All-Day Recordings and Automated Analysis. Am J Speech Lang Pathol. 2017;26(2):248–265. doi:10.1044/2016_AJSLP-15-0169

20. Bang JY, Kachergis G, Weisleder A, Marchman V. An automated classifier for periods of sleep and target-child-directed speech from LENA recordings. Lang Dev Res. 2023;3(1). doi:10.34842/xmrq-er43

21. Bates D, Mächler M, Bolker B, Walker S. Fitting Linear Mixed-Effects Models Using lme4. J Stat Softw. 2015;67:1–48. doi:10.18637/jss.v067.i01

22. Byrne EM, Hunt K, Scala M. Introducing the i-Rainbow©: An Evidence-Based, Parent-Friendly Care Pathway Designed for Critically Ill Infants in the NICU Setting. Pediatr Phys Ther. 2024;36(2):266. doi:10.1097/PEP.0000000000001094

